# REAL-WORLD IMPACT OF THE FIRST WAVE OF THE COVID-19 PANDEMIC ON THE CYSTIC FIBROSIS COMMUNITY THROUGHOUT EUROPE

**DOI:** 10.1101/2025.04.11.25325660

**Authors:** Emmanuelle Bardin, Elise Lammertyn, Hilde De Keyser

## Abstract

**Background:** During the outbreak of the COVID-19 pandemic in the Spring of 2020, lockdowns were imposed in an attempt to stop its spreading. This situation was particularly concerning for people living with a chronic condition, such as people with cystic fibrosis (PWCF), who are vulnerable to respiratory infection and need regular clinical follow-ups.

**Methods:** In April 2020, a Europe-wide survey investigated the impact of the pandemic-related restrictions on the day-to-day life, care and wellbeing of people living with a rare disease. Responses of the CF community are presented with a focus on the differential impact between Eastern European countries (EEC) and Western European countries (WEC).

**Results:** Access to hospitals and professional care was greatly reduced, and more than 60% of respondents felt isolated, depressed or helpless. This fed the perception that this crisis was detrimental to health and 92% of respondents to consider COVID-19 as high threat to PWCF. Although mortality rate was limited in EEC during this first wave, shortages and cancellations were reported twice as often compared to WEC, and EEC respondents were more fearful to visit the hospital. Incidentally, the survey revealed a large ‘employment gap’ between EEC and WEC, generating financial insecurity in EEC and higher apprehension.

**Conclusions:** This peril had a major impact on psychosocial wellbeing and highlighted the fact that PWCF relied on the community support. On the other hand, it triggered the integration of telemedicine into routine CF care. European umbrella organisations must aim to coordinate efforts and harmonise access to care, treatments and support throughout Europe.

**Declarations of interest:** none

## Introduction^1^

The World Health Organisation (WHO) first reported on a novel coronavirus causing outbreaks of pneumonia of unknown aetiology in (South)east Asia in January 2020 and declared the ensuing coronavirus disease 19 (COVID-19) a pandemic on 11 March 2020 (1). To stop SARS-CoV-2 virus from spreading, many countries took precautionary measures such as closing schools, imposing so-called social distancing and lockdown.

Cystic fibrosis (CF) is one of the most prevalent autosomal recessive disorders worldwide with more than 49 000 patients known in Europe (2). The 2009 H1N1 influenza pandemic resulted in significant morbidity in people with CF (PWCF) (3). Therefore, PWCF were considered highly vulnerable to SARS-CoV-2, with an increased risk of complications, recommending self-isolation or “shielding” and reinforcing the preventing measures already well-established in this population, e.g. face masks and hand hygiene. During the first pandemic wave in the spring of 2020, 130 PWCF contracted SARS-CoV- 2 in Europe and incidence amongst the CF population was higher than in the age-matched general population (4). Most patients experienced mild symptoms, 58.1% were hospitalised amongst whom 9.2% needed intensive care, and 5 died (5). Whilst outcomes may be better than initially feared, PWCF suffer from more severe clinical course (6), which is possibly associated with transplantation, older age, CF-related diabetes and lower lung function (7).

During the first COVID-19 wave, Western and Northern countries were severely affected and suffered a high mortality rate despite a higher health service capacity, whilst Eastern countries with a weaker healthcare system reacted swiftly and implemented containment measures at the same time, which limited mortality rate (8–11). Many societal studies have described how countries throughout the globe coped with the first pandemic wave. Amongst the CF community, the pandemic resulted in changes and cancellation of routine medical appointments (12); an early Irish survey reported concerns about the ability to access hospital care (13). Particular attention was paid to the consequences of the pandemic and precautionary measures on mental wellbeing. Various national surveys report evident psychological impact with respondents experiencing increased stress, fear and anxiety, both amongst people living with a chronic and/or rare disease (14–16), including CF (12,13,17–19), and in the general population (20–24).

EURORDIS is a non-governmental patient-driven alliance of patient organisations (POs) representing 956 rare disease POs in 73 countries. From 18 April to 11 May 2020, EURORDIS conducted a large-scale survey investigating the impact of COVID-19 on the life and care of people living with a rare disease or their carers in Europe. Here we present the responses of PWCF and their relatives participating in the EURORDIS survey, bringing insight into the consequences of the first pandemic wave on the day-to- day life of the CF community with a focus on the differential impact between lower income Eastern European countries (EEC) and wealthier Western European countries (WEC).

## Methods

The electronic survey (SphinxOnline, Annecy, France) was conducted in the framework of EURORDIS Rare Barometer Programme and disseminated by Cystic Fibrosis Europe (CFE), the federation of national CF POs from 39 European countries. The survey was available in 23 languages and included 23 questions of which 16 were relevant for CF, categorised into five themes (Table 1, answers in Table S1).

**Table 1.**
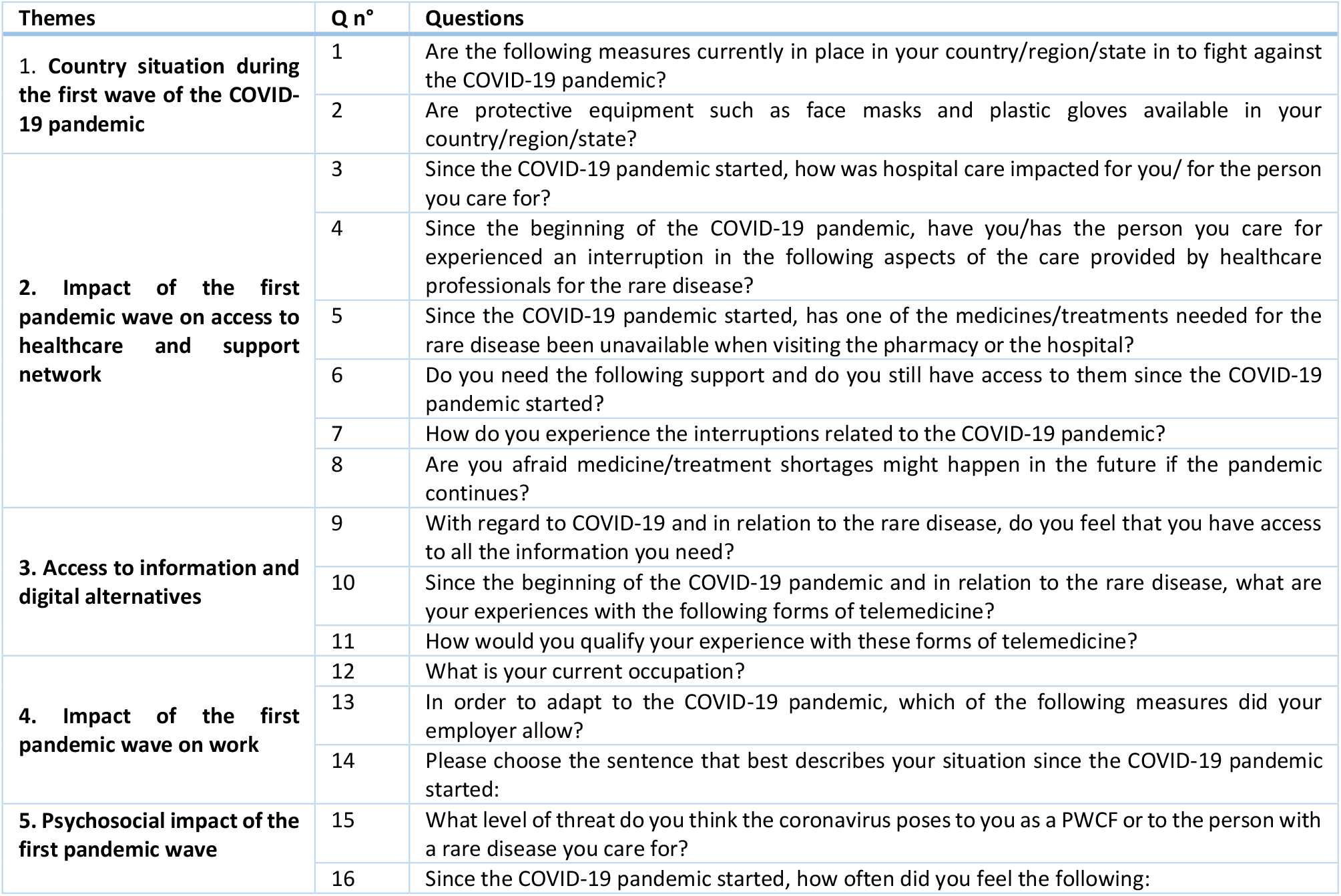
List of the questions in the survey.

| Themes | Q n° | Questions |
| --- | --- | --- |
| **1. Country situation during the first wave of the COVID-19 pandemic** | 1 | Are the following measures currently in place in your country/region/state in to fight against the COVID-19 pandemic? |
|  | 2 | Are protective equipment such as face masks and plastic gloves available in your country/region/state? |
| **2. Impact of the first pandemic wave on access to healthcare and support network** | 3 | Since the COVID-19 pandemic started, how was hospital care impacted for you/ for the person you care for? |
|  | 4 | Since the beginning of the COVID-19 pandemic, have you/has the person you care for experienced an interruption in the following aspects of the care provided by healthcare professionals for the rare disease? |
|  | 5 | Since the COVID-19 pandemic started, has one of the medicines/treatments needed for the rare disease been unavailable when visiting the pharmacy or the hospital? |
|  | 6 | Do you need the following support and do you still have access to them since the COVID-19 pandemic started? |
|  | 7 | How do you experience the interruptions related to the COVID-19 pandemic? |
|  | 8 | Are you afraid medicine/treatment shortages might happen in the future if the pandemic continues? |
| **3. Access to information and digital alternatives** | 9 | With regard to COVID-19 and in relation to the rare disease, do you feel that you have access to all the information you need? |
|  | 10 | Since the beginning of the COVID-19 pandemic and in relation to the rare disease, what are your experiences with the following forms of telemedicine? |
|  | 11 | How would you qualify your experience with these forms of telemedicine? |
| **4. Impact of the first pandemic wave on work** | 12 | What is your current occupation? |
|  | 13 | In order to adapt to the COVID-19 pandemic, which of the following measures did your employer allow? |
|  | 14 | Please choose the sentence that best describes your situation since the COVID-19 pandemic started: |
| **5. Psychosocial impact of the first pandemic wave** | 15 | What level of threat do you think the coronavirus poses to you as a PWCF or to the person with a rare disease you care for? |
|  | 16 | Since the COVID-19 pandemic started, how often did you feel the following: |

Respondents were classified as either from EEC or from WEC (Table S2). Geographic disparities and the low number of responses from most individual countries precluded from applying a relevant statistical correction. The weight of France and Greece, which had the highest numbers of respondents in WEC and EEC respectively, was evaluated by excluding each of them from data processing. Although distributions were impacted, these countries did not drive alone the overall trends and conclusions, indicating robust and consistent data. Thus, the raw data are presented, after normalisation to the total number of respondents to each question. Exhaustive data are available in Table S3.

Correlation coefficients were calculated to assess possible links between, on one hand, variables explored in Q1, Q2, Q4, Q5, Q6, Q7, Q8 and Q9, and on the other hand, psychological status of respondents (Q15 and Q16). These being qualitative ordered data, the Kendall tau method was employed. Answers that could be ordered were assigned a quantitative value, others were excluded (Table S4). Correlations were considered significant when their associated *p*-value was below 1,9.10^−4^, thereby accounting for a risk of 5% and for multiple testing using the Bonferroni correction.

## Results

### Population characteristics

There were 897 respondents from 29 countries (Table S5, Figure S1); 81% lived in WEC and 78% were females. Comparable numbers of PWCF and carers took the survey (52% and 48%). Most of the latter were parents, 86% being women. More PWCF filled out the survey in WEC than in EEC (54% vs. 43%). Age distribution was similar in WEC and EEC. Amongst PWCF, the age category below 35 was overrepresented (59%), whereas most of the other respondents were 35-49y (52%). In the “50 and above” age category, the number of carers was twice as high as the number of PWCF.

### Theme 1: Country situation during the first wave of COVID-19 pandemic

Social distancing measures and closure of educational facilities were comparable in EEC and WEC (92%- 98% of respondents) (Q1). Yet, lockdown measures were more prevalent in WEC (82% vs. 72% in EEC). Respondents reported that personal protective equipment (PPE) was easier to find in EEC than in WEC (44% vs. 13%) (Q2, Figure S2). About 7% of the WEC respondents said they did not need PPE.

### Theme 2: Impact on access to healthcare and support

More than 75% of EEC respondents skipped their hospital appointment out of fear of COVID-19 whereas they were about 60% in WEC. In parallel, 30% of respondents in EEC and 15% in WEC reported closed hospital units. Equipment usually allocated to PWCF care was reported missing by 10% in EEC and 16% in WEC.

The proportion of postponements and cancellations taken together varied from 40% to 80% in all aspects of CF health care. Rehabilitation therapies, e.g. respiratory physiotherapy, were the most impacted with about 80% delays or cancellations in both WEC and EEC. Other appointments were similarly impacted in WEC with an average of 60% cancellation/postponement. In EEC, there were twice more cancellations in follow-ups and diagnostic tests and a total above 75% cancellation/postponement. Surgeries and psychological follow-ups were equally cancelled in WEC and EEC (15%) but there were more delays in WEC.

Although complete unavailability was rare, temporary medicine shortages were reported, notably more frequently in EEC (Q5, Table S3).

According to respondents, impact on support availability was limited (Q6; Figure 1c). The most important support was family and social network, which remained mostly accessible. Professional support deemed inaccessible in EEC was usually already so before the pandemic though the need seemed higher there than in WEC.

**Figure 1.**
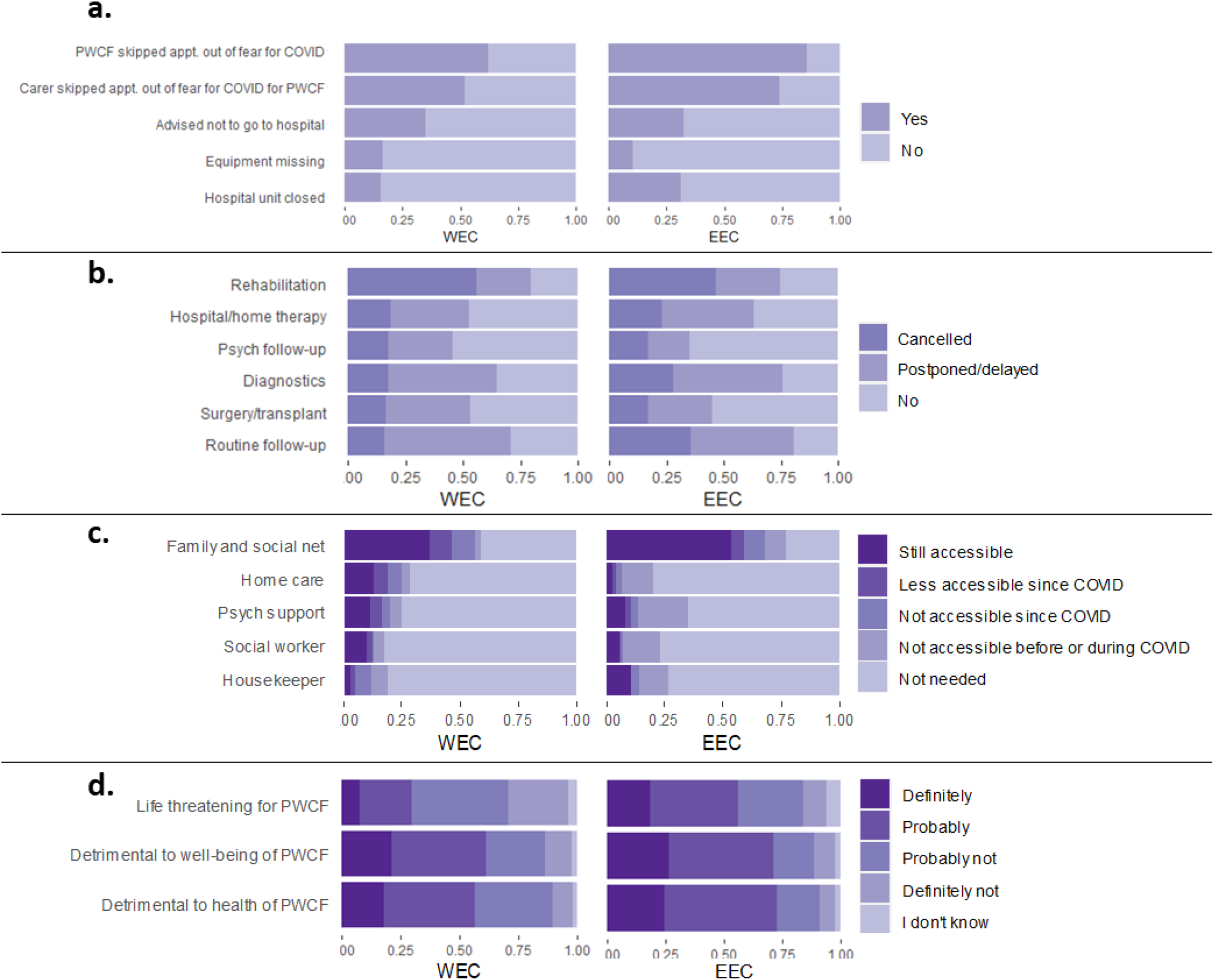
Impact on healthcare and support network. Distribution expressed as percentages of the total number of responses to **a**. Q3: Since the COVID-19 pandemic started, how was hospital care impacted for you/for the person you care for? **b**. Q4: Since the beginning of the COVID-19 pandemic, have you/has the person you care for experienced an interruption in the following aspects of the care provided by healthcare professionals for the rare disease? **c**. Q6: Do you need the following support and do you still have access to them since the COVID-19 pandemic started? **d**. Q7: How do you experience the interruptions related to the COVID-19 pandemic?.

Interruptions in healthcare were perceived as detrimental to the wellbeing and health of PWCF by ∼60% of respondents in WEC and 75% in EEC, and life-threatening by 25% in WEC and 50% in EEC (Q7; Figure 1d). PWCF seemed more preoccupied than their carers (data not shown). Respondents below 35 and above 50 appeared more pessimistic than the 35-49 age group (data not shown).

Overall, 85% of respondents perceived the future with apprehension when it came to the availability of treatments (Q8, Table S3).

### 1. Theme 3: Access to information and alternative resources

Across all age ranges, about 25% of WEC respondents felt a lack of information (“never” or “seldom” access to information needed), compared to 15% in EEC (Q9; Figure 2a). Interestingly, the younger population was more in demand compared to respondents above 50.

**Figure 2.**
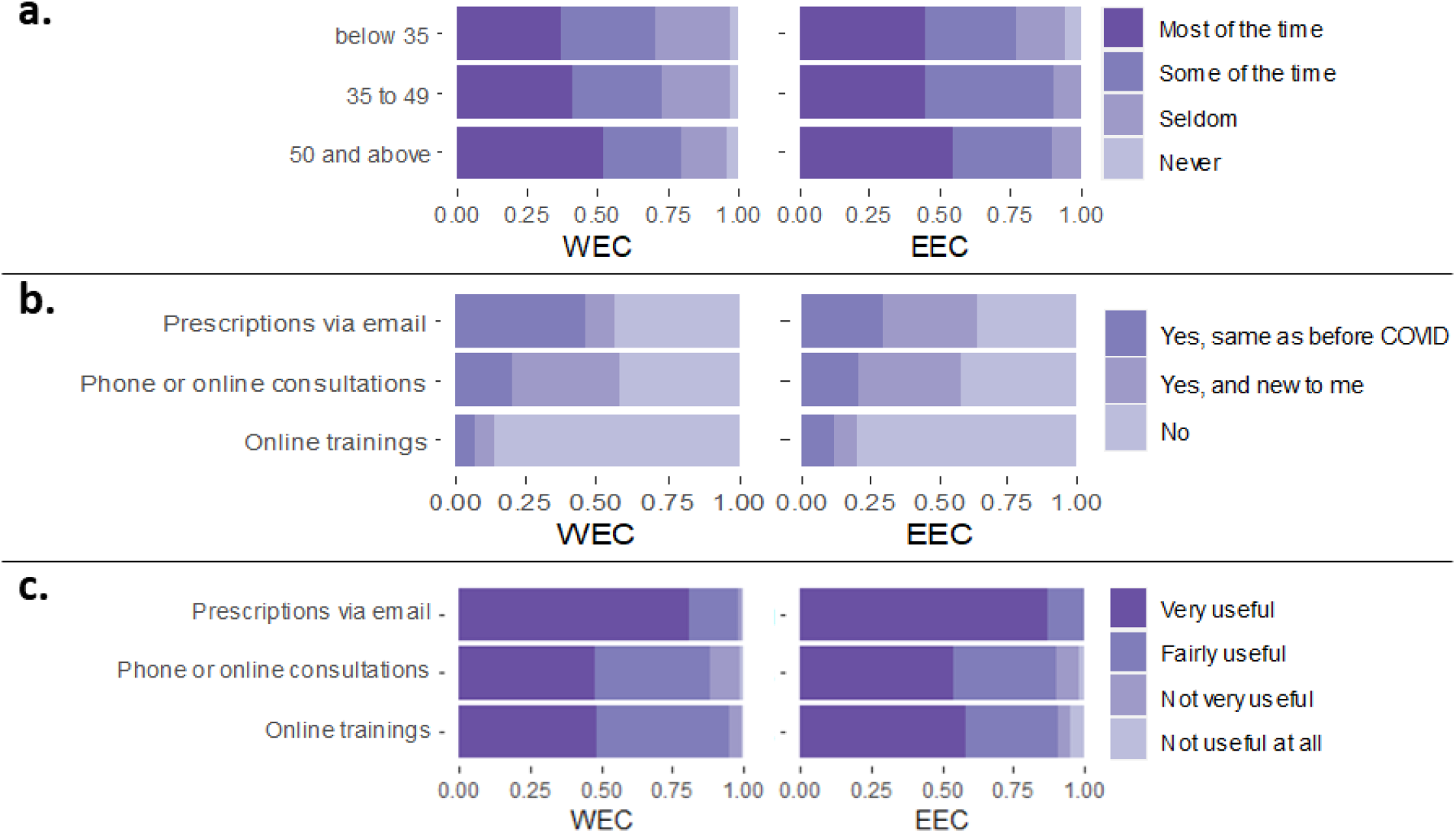
Access to information and digital alternatives. Distribution expressed as percentages of the total number of responses to **a**. Q9: With regard to COVID-19 and in relation to the rare disease, do you feel that you have access to all the information you need? **b**. Q10: Since the beginning of the COVID-19 pandemic and in relation to the rare disease, what are your experiences with the following forms of telemedicine? **c**. Q11: How would you qualify your experience with these forms of telemedicine?

In Q10, 36% to 44% of total respondents indicated not to use phone/online consultations or email prescriptions (Figure 2b). A total of 58% respondents received email prescriptions during the pandemic, levelling the pre-existing gap between EEC and WEC (30% vs 46%, respectively, before the pandemic). Online consultations increased with the emergence of the pandemic; yet only 15% of respondents had access to online training.

Telemedicine was globally perceived as useful, especially email prescriptions (Q11; Figure 2c).

### 2. Theme 4: Impact on work

Half of respondents declared to be employed but there were considerable differences between WEC (54%) and EEC (32%) (Q12; Figure 3a). In EEC, more respondents were unemployed, retired or on leave of absence. 42% of PWCF and 58% of carers were employed (data not shown). Logically, the patient population included more people “unable to work” (21%) and students (14%) than the carer population (2% and 3%, respectively). There were more homemakers amongst relatives (14%) than amongst PWCF.

**Figure 3.**
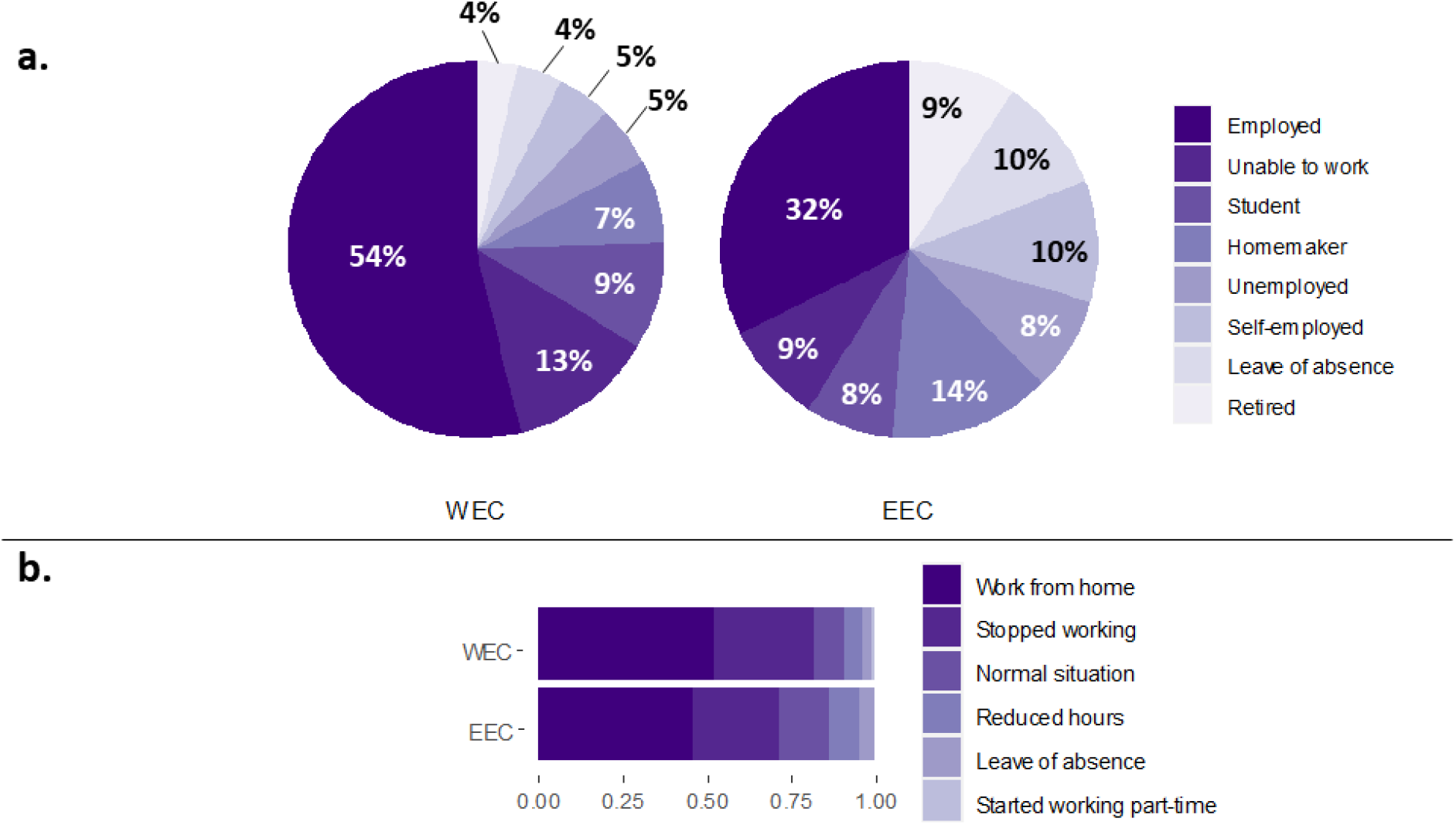
Impact on work. Distribution expressed as percentages of the total number of responses to **a**. Q12: What is your current occupation? **b**. Q14: Please choose the sentence that best describes your situation since the COVID-19 pandemic started.

Although half of the total respondents to Q14 stated to work from home since the beginning of the pandemic (Figure 3b), more than 25% were not given this opportunity (Q13; Table S3) and more than a quarter had to stop working. About 10% of respondents stated the pandemic had no impact on their working modalities with a higher proportion in EEC.

### 3. Theme 5: Psychosocial impact

EEC appeared systematically slightly more impacted (Figure 4). The pandemic was perceived as a high to very high threat to PWCF by a large majority (91% in WEC, 94% in EEC) (Figure 4a).

**Figure 4.**
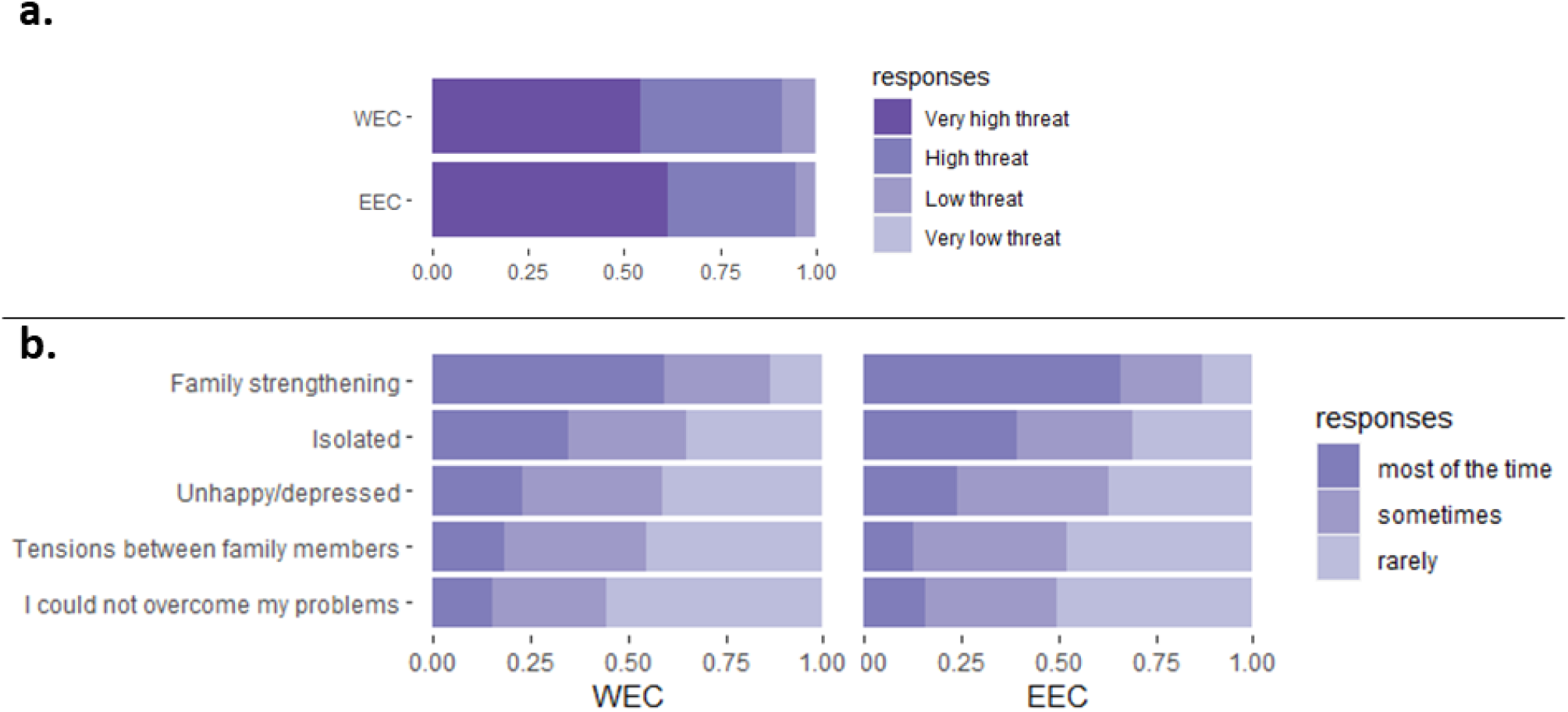
Psychosocial impact. Distribution expressed as percentages of the total number of responses to **a**. Q15: What level of threat do you think the coronavirus poses to you as a PWCF or to the person with a rare disease you care for? **b**. Q16: Since the COVID-19 pandemic started, how often did you feel the following.

Two thirds of respondents felt isolated (65% in WEC, 69% in EEC) or depressed (59% in WEC, 65% in EEC) at some point and almost half faced difficulties to overcome problems (44% in WEC, 50% in EEC) (Figure 4b). Although half of respondents experienced tensions between family members, 87% also reported family strengthening.

### 4. Impact of pandemic-related factors on psychological status

Since trends were similar between WEC and EEC regarding psychological impact, correlations were calculated on pooled data. There were no significant correlations with country-specific situations (Q1 and Q2). Most of significant coefficients were around +/- 0,25 (Table S6). The strongest correlations were found between “feeling depressed” and “difficulties to overcome problems” (+ 0,56) or “feeling isolated” (+0,46). “Feeling depressed” was further correlated with “family tensions” (+ 0,33) and the perception that interruptions in healthcare were detrimental to wellbeing (+ 0,24).

“Isolation” was also correlated to “difficulties to overcome problems” (+ 0,38) and “family tensions” (+ 0,23).

“Difficulties to overcome problems” further correlated with “family tensions” (+0,27) and the whole range of healthcare interruptions. It also weakly correlated with a lack of information (−0,14).

“Family strengthening” negatively correlated with “feeling depressed” (− 0,17) and “feeling isolated” (− 0,15).

The level of threat felt by PWCF correlated with the perception of healthcare interruptions being detrimental to health (+ 0,26) or life-threatening (+ 0,28). Notably, these correlations were higher in carers’ responses (+ 0,30) and the level of threat felt for the person cared for was also linked to the fear of shortages (+ 0,25).

## Discussion

### Population characteristics and limitations

With about 900 participants, this is to our knowledge the largest real-life study exploring the impact of the first COVID-19 wave on the CF community throughout Europe both practically and psychologically, incidentally bringing insight into the pre-pandemic situation, notably regarding support availability and work situation. Its main limitation resides in the disparity of the respondent population, in terms of age, gender and geography, which introduces bias to keep in mind.

The shorter life expectancy of PWCF and the fact that most of carers were parents resulted in a younger population of PWCF respondents. As is common in such surveys, there was a higher proportion of females; mothers represented more than a third of the total respondents. There is thus a discrepancy between how mothers and young adults with CF may handle and fear the pandemic consequences.

Importantly, there were four times more respondents from WEC than EEC. Possible reasons are a larger (known) CF population in WEC, more efficient dissemination, more awareness and more exposition to the virus at the time as well as easier access to IT tools in WEC. However, these do not explain the imbalance observed at the country level within WEC and within EEC. For example, respondents from the Netherlands and Greece were 10 times more numerous than those from the United Kingdom (UK), despite its larger CF population. This may reflect ‘survey saturation or fatigue’, due to other on-going national surveys, and variable levels of engagement from patient representatives in disseminating this questionnaire.

Noteworthy, we investigated the respondents’ perspective, whose subjective perception may differ from the reality. Furthermore, this kind of surveys more easily reaches people already active, especially online, and engaged in patient advocacy.

Finally, whilst we intended to compare the impact of the first wave in WEC and EEC, confounding factors concur and may not always be discernible, e.g. EEC facing a lower SARS-CoV-2 circulation than WEC at the time.

### Differential impact on EEC and WEC

EEC managed to better limit mortality rate during the first episode of the pandemic by introducing prevention measures at the same time as WEC. Nevertheless, and even though PPE seemed still accessible in EEC contrary to WEC, EEC respondents were more fearful to visit the hospital. Moreover, closed hospitals, temporary shortages, cancelled appointments were reported twice as often in EEC compared to WEC. In addition to the obvious risks faced by PWCF due to disrupted care, these observations point towards a pre-existing, less resilient healthcare provision for PWCF, in contrast with more integrated CF care in WEC (9). This is also illustrated by the lack of professional support dating back from before the pandemic in EEC.

Furthermore, although there were more people working normally in EEC, which is consistent with less prevalent lockdown measures, the data mostly show a large ‘employment gap’ between WEC and EEC, where only one third of participants replied to be employed. The proportion of homemakers was twice as high in EEC compared to WEC, which can be attributed at least partially to a shortfall in integrated care and support for people living with a chronic condition. These elements suggest a lower income and lower overall socioeconomic status for PWCF and their families, which is known to affect clinical and patient-reported outcomes (25). Although far from ideal in WEC, the integration of people with health limitations into the labour market seems more advanced than in EEC.

These practicalities negatively impacted the capacity of the CF community to cope with the pandemic in EEC, resulting in a much higher apprehension and an increased perception of life threat compared to their WEC peers. Even though EEC were praised for their successful initial containment of SARS-CoV- 2, these countries were devastatingly struck by the second wave in Autumn 2020, legitimating the apprehension of EEC respondents revealed by this survey.

### Access to information and digital alternatives to face the pandemic

Information about SARS-CoV-2 was scarce and inconsistent at the beginning of the pandemic, when people were clearly eager to know more about the risks and possible consequences of an infection. Vulnerable populations especially needed continuous personalised recommendations (26,27). In the present study, 10% to 30% of respondents felt they seldom or never had access to the needed information. The feeling prevailed amongst the western population, more exposed to the virus, and individuals below 35, who are used to an easy and wide access to information. This shortfall of information led to a poor visibility on the future and a feeling of helplessness, i.e. difficulty to overcome problems.

On a brighter side, access to telemedicine was important and likely played a substantial role in ensuring some continuity of care for PWCF (28). Accordingly, a Turkish study showed that patients with a chronic lung disease found these tools more useful than the general population (14). Furthermore, the pandemic seems to have accelerated use, access and diversity of innovative solutions in general, enabling EEC to reach a level comparable to WEC, although respondents to such online surveys are already familiar with digital tools.

Such approaches could be advantageous for PWCF and patients on the long-term, as long as they are fit for purpose and do improve care. Remote consultations may be appropriate to discuss results of a routine medical analyses or to adjust a prescription (29). Access to mental health services through telemedicine has proven to be effective in the context of depression and might be useful in reducing the mental health burden in populations practicing long-term shielding (30). However, when delicate topics are to be discussed, e.g. transplantation or maternity, face-to-face consultations are preferred. Also, technical examinations, such as lung functional tests or paediatric follow-ups are preferably done at the hospital. Furthermore, one must be aware of a ‘remote medicine bias’: patients might declare to be fine to avoid a visit to the hospital. Moving too rapidly towards telemedicine as a permanent model for care delivery could undo some of the progress achieved over recent years, such as trustful relationships with the medical profession and treatment compliance. Furthermore, it could widen the CF care gap based on economic resources as it creates a dependence on access to a smartphone or high-speed internet (31).

### Psychological impact and the key role of community support

There were no statistically significant correlations between psychological status and country-specific measures or situations, though isolation and care disruption had a clear impact on mental health. Together with family tensions, they generated an overall impression of helplessness which translated into a feeling of depression for two thirds of respondents. People particularly worried about the impact of healthcare disruptions resulting in more than 90% respondent considering the situation as a high to very high threat. This confirms other national studies’ findings, all observing an increased degree of anxiety since the beginning of the pandemic in PWCF (12,13,18,19) or in people with a chronic respiratory disease (14). Surveys on other chronic diseases such as hemoglobinopathies also reported a clear link between care disruptions, isolation and deterioration of mental health (32,33).

Anxiety manifested stronger in the younger and older respondents than the intermediate age group, a phenomenon that has also been observed in the healthy population (22–24). The younger may feel anxious of the long-term consequences of such interruptions, whereas the older population may feel more isolated and vulnerable to COVID-19.

Of note, some studies reported no significant links between a pre-existing condition and a higher level of anxiety at the time (34,35). According to an Italian survey, PWCF even tended to have lower psychological distress than the general population when coping with the lockdown (36). This may be thanks to more mature coping styles amongst patients as described by a Turkish study, though disease was also associated with higher anxiety (14). Another Italian study on thalassemia depicts a similar trend, with higher levels of anxiety but transcendent coping strategies compared to controls (37).

Furthermore, despite the pandemic, a quarter of respondents were not given more flexibility at work and another quarter had to stop working, which may have generated concerns regarding both short and long-term economic consequences. This stresses the necessity to encourage employers to allow their employees, especially those at higher risk, to work flexibly and/or from home when possible. This should increase the feeling of safety, trust and wellbeing at work, also beyond this pandemic.

Feelings were also correlated with family relationships. Tensions surfaced, maybe due to the unusual proximity imposed by lockdown and general anxiety; yet family strengthening was the most important support against isolation and depression. Strikingly, most of the respondents declared not to need the other listed supports, emphasizing the importance of family and friends supports in people living with a chronic disease (29). Altogether, it demonstrates how resilient PWCF are and how strong the CF community stands (14,36).

### There is a need for a European umbrella guidance

This survey highlighted differences between EEC and WEC in terms of practicalities, such as access to support and employment, but also in terms of organisation and empowerment. Indeed, it is more difficult for the smaller number of PWCF living in an EEC to get their voice heard, compared to WEC with well-established POs. Hence, there is a need for umbrella organisations, such as CFE and EURORDIS, to provide a Europe-wide overview and to advocate for patients living with a rare disease such as CF in more challenging conditions. Such an organisation must aim to coordinate the efforts of WEC and EEC and to harmonise access to care, treatments and support. Following the EURORDIS survey, Castro et al listed a number of recommendations aiming to ensure to the most vulnerable populations throughout Europe effective integrated and holistic care pathways (29).

## Data Availability

The data underlying this study were collected anonymously via an online survey and are not publicly available due to privacy considerations and the terms of participant consent. Data may be made available by the authors upon reasonable request and with permission from EURORDIS - Rare Diseases Europe.

## ^1^ Abbreviations

COVID-19: coronavirus disease 19
PWCF: people with cystic fibrosis
EEC: Eastern European countries
WEC: Western European countries
WHO: World Health Organisation
PO: patient organisation
PPE: personal protective equipment.

## Author contributions

EB: conceptualization, data curation, formal analysis, investigation, methodology, visualisation, writing – original draft, writing – review & editing; EL: formal analysis, project administration, writing – original draft, writing – review & editing; HDK: conceptualization, supervision, writing – review and editing

## Acknowledgements

We wish to thank our colleagues from EURORDIS and the Rare Barometer for designing the survey and sharing the collected data as well as the local and national POs and patient representatives who helped disseminating the survey. We are grateful to Maya Kirszenbaum (Hôpital Necker Enfants Malades,

Paris, France) for her helpful insights and we are very thankful to all people living with cystic fibrosis and their carers who took the survey.

## References

1. World Health Organization. Coronavirus Disease (COVID-19) Situation Reports [Internet]. 2020 [cited 2020 Oct 13]. Available from: https://www.who.int/emergencies/diseases/novel-coronavirus-2019/situation-reports

2. Orenti A, Zolin A, Van Rens J, Fox A, Krasnyk M, Mei-Zahav M, et al. ECFS Patient Registry 2020. ECFS; 2022 p. 163.

3. Viviani L, Assael BM, Kerem E. Impact of the A (H1N1) pandemic influenza (season 2009–2010) on patients with cystic fibrosis. J Cyst Fibros. 2011 Sep 1;10(5):370–6.

4. Naehrlich L, Orenti A, Dunlevy F, Kasmi I, Harutyunyan S, Pfleger A, et al. Incidence of SARS-CoV-2 in people with cystic fibrosis in Europe between February and June 2020. J Cyst Fibros. 2021 Jul;20(4):566–77.

5. European Cystic Fibrosis Society Patient Registry. COVID-CF project in Europe [Internet]. [cited 2021 Aug 31]. Available from: https://www.ecfs.eu/covid-cf-project-europe

6. Jung A, Orenti A, Dunlevy F, Aleksejeva E, Bakkeheim E, Bobrovnichy V, et al. Factors for severe outcomes following SARS-CoV-2 infection in people with cystic fibrosis in Europe. ERJ Open Res. 2021 Oct;7(4):00411–2021.

7. Carr SB, McClenaghan E, Elbert A, Faro A, Cosgriff R, Abdrakhmanov O, et al. Factors associated with clinical progression to severe COVID-19 in people with cystic fibrosis: A global observational study. J Cyst Fibros Off J Eur Cyst Fibros Soc. 2022 Jul;21(4):e221–31.

8. Ding X, Cai Z, Zhu W, Fu Z. Study on the Spatial Differentiation of Public Health Service Capabilities of European Union under the Background of the COVID-19 Crisis. Healthcare. 2020 Sep 24;8(4):358.

9. Petrović D, Petrović M, Bojković N, Čokić VP. An integrated view on society readiness and initial reaction to COVID–19: A study across European countries. Adrish M, editor. PLOS ONE. 2020 Nov 23;15(11):e0242838.

10. James N, Menzies M, Radchenko P. COVID-19 second wave mortality in Europe and the United States. Chaos Interdiscip J Nonlinear Sci. 2021 Mar;31(3):031105.

11. Lupu D, Tiganasu R. COVID-19 and the efficiency of health systems in Europe. Health Econ Rev. 2022 Dec;12(1):14.

12. Havermans T, Houben J, Vermeulen F, Boon M, Proesmans M, Lorent N, et al. The impact of the COVID-19 pandemic on the emotional well-being and home treatment of Belgian patients with cystic fibrosis, including transplanted patients and paediatric patients. J Cyst Fibros. 2020 Nov;19(6):880–7.

13. Ring E. Half of people with cystic fibrosis fear contracting Covid-19 [Internet]. Irish Examiner. 2020 [cited 2021 Aug 31]. Available from: https://www.irishexaminer.com/news/arid-30993128.html

14. Ademhan Tural D, Emiralioglu N, Tural Hesapcioglu S, Karahan S, Ozsezen B, Sunman B, et al. Psychiatric and general health effects of COVID-19 pandemic on children with chronic lung disease and parents’ coping styles. Pediatr Pulmonol. 2020 Dec;55(12):3579–86.

15. Sloan M, Gordon C, Lever E, Harwood R, Bosley MA, Pilling M, et al. COVID-19 and shielding: experiences of UK patients with lupus and related diseases. Rheumatol Adv Pract. 2021 Jan 25;5(1):rkab003.

16. EURORDIS. How has COVID-19 impacted people [Internet]. 2020. Available from: https://www.eurordis.org/publications/how-has-covid-19-impacted-people-with-rare-diseases/

17. Pınar Senkalfa B, Sismanlar Eyuboglu T, Aslan AT, Ramaslı Gursoy T, Soysal AS, Yapar D, et al. Effect of the COVID-19 pandemic on anxiety among children with cystic fibrosis and their mothers. Pediatr Pulmonol. 2020 Aug;55(8):2128–34.

18. Rhoads S, Cooney K, Banerjee D. Emotional Impact of COVID-19 Pandemic on Adults with Cystic Fibrosis. C O M M E N TA R. :3.

19. Westcott KA, Wilkins F, Chancellor A, Anderson A, Doe S, Echevarria C, et al. The impact of COVID-19 shielding on the wellbeing, mental health and treatment adherence of adults with cystic fibrosis. Future Healthc J. 2021 Mar;8(1):e47–9.

20. Buzzi C, Tucci M, Ciprandi R, Brambilla I, Caimmi S, Ciprandi G, et al. The psycho-social effects of COVID-19 on Italian adolescents’ attitudes and behaviors. Ital J Pediatr. 2020 Dec;46(1):69.

21. Dawel A, Shou Y, Smithson M, Cherbuin N, Banfield M, Calear AL, et al. The Effect of COVID-19 on Mental Health and Wellbeing in a Representative Sample of Australian Adults. Front Psychiatry. 2020 Oct 6;11:579985.

22. Toffolutti V, Plach S, Maksimovic T, Piccitto G, Mascherini M, Mencarini L, et al. The association between COVID-19 policy responses and mental well-being: Evidence from 28 European countries. Soc Sci Med. 2022 May;301:114906.

23. Di Gessa G, Price D. The impact of shielding during the COVID-19 pandemic on mental health: evidence from the English Longitudinal Study of Ageing. Br J Psychiatry. 2022 Apr 4;1–7.

24. Hyland P, Shevlin M, McBride O, Murphy J, Karatzias T, Bentall RP, et al. Anxiety and depression in the Republic of Ireland during the COVID-19 pandemic. Acta Psychiatr Scand. 2020 Sep;142(3):249–56.

25. Quittner AL, Schechter MS, Rasouliyan L, Haselkorn T, Pasta DJ, Wagener JS. Impact of Socioeconomic Status, Race, and Ethnicity on Quality of Life in Patients With Cystic Fibrosis in the United States. CHEST. 2010 Mar 1;137(3):642–50.

26. Collaço N, Legg J, Day M, Culliford D, Campion A, West C, et al. COVID-19: Impact, experiences, and support needs of children and young adults with cystic fibrosis and parents. Pediatr Pulmonol. 2021 Sep;56(9):2845–53.

27. Banchev A, Batorova A, Kotnik BF, Kiss C, Puras G, Zapotocka E, et al. A Cross-National Survey of People Living with Hemophilia: Impact on Daily Living and Patient Education in Central Europe. Patient Prefer Adherence. 2021 Apr 28;15:871–83.

28. Smith BA, Georgiopoulos AM, Mueller A, Abbott J, Lomas P, Aliaj E, et al. Impact of COVID-19 on mental health: Effects on screening, care delivery, and people with cystic fibrosis. J Cyst Fibros. 2021 Dec;20:31–8.

29. Castro R, Berjonneau E, Courbier S. Learning from the Pandemic to Improve Care for Vulnerable Communities: The Perspectives and Recommendations from the Rare Disease Community. Int J Integr Care. 2021 Mar 4;21(1):12.

30. Zhou X, Snoswell CL, Harding LE, Bambling M, Edirippulige S, Bai X, et al. The Role of Telehealth in Reducing the Mental Health Burden from COVID-19. Telemed E-Health. 2020 Apr 1;26(4):377–9.

31. Davies J. The coronavirus pandemic has forced rapid changes in care protocols for cystic fibrosis. Nature. 2020 Jul 29;583(7818):S15–S15.

32. Delicou S, Xydaki A, Manganas K, Koullias E, Evliati L, Kalkana C, et al. The Effect of COVID-19 on Hemoglobinopathy Patients’ Daily Lives While Quarantined: Four Greek Hospitals’ Experiences. Thalass Rep. 2022 Jun;12(2):39–45.

33. Arian M, Vaismoradi M, Badiee Z, Soleimani M. Understanding the impact of COVID-19 pandemic on health-related quality of life amongst Iranian patients with beta thalassemia major: a grounded theory. Prim Health Care Res Dev. 2021 ed;22:e67.

34. Budu MO, Rugel EJ, Nocos R, Teo K, Rangarajan S, Lear SA. Psychological Impact of COVID-19 on People with Pre-Existing Chronic Disease. Int J Environ Res Public Health. 2021 Jun 2;18(11):5972.

35. Louvardi M, Pelekasis P, Chrousos GP, Darviri C. Mental health in chronic disease patients during the COVID-19 quarantine in Greece. Palliat Support Care. 2020 Aug;18(4):394–9.

36. Ciprandi R, Bonati M, Campi R, Pescini R, Castellani C. Psychological distress in adults with and without cystic fibrosis during the COVID-19 lockdown. J Cyst Fibros. 2021 Mar;20(2):198–204.

37. Cerami C, Santi GC, Sammartano I, Borsellino Z, Cuccia L, Battista Ruffo G, et al. Uncertain crisis time affects psychosocial dimensions in beta-thalassemia patients during Covid-19 pandemic: A cross-sectional study. J Health Psychol. 2022 Sep 1;27(11):2529–38.

