## Supplemental Tables and Figures for "REAL-WORLD IMPACT OF THE FIRST WAVE OF THE COVID-19 PANDEMIC ON THE CYSTIC FIBROSIS COMMUNITY THROUGHOUT EUROPE"

*Table S1. List of questions and related options and responses*

| Q | Question | Options | Responses |
|---|---|---|---|
| **1** | Are the following measures currently in place in your country/region/state in to fight against the COVID-19 pandemic? | a) Social distancing, which includes avoiding mass gatherings, and maintaining distance from others when possible<br>b) Closure of educational facilities<br>c) Lockdown measures | - Yes<br>- No<br>- I don't know |
| **2** | Are protective equipment such as face masks and plastic gloves available in your country/region/state? | a) For you<br>b) For the person you care for<br>c) For your health professionals<br>d) For your social care professionals | - Available and easily accessible<br>- Difficult to find, sometimes not available<br>- Impossible to find, not available<br>- Not needed |
| **3** | Since the COVID-19 pandemic started, how was hospital care impacted for you/ for the person you care for? | a) You did not go to the hospital because you are fearful of catching COVID-19<br>b) You did not go to the hospital because you are fearful that the person you care for might catch COVID-19<br>c) Being told not to go to the hospital if you or the person you care for affected by a rare disease becomes unwell for other reasons than COVID-19<br>d) The hospital/unit that provides care for the rare disease is closed<br>e) The necessary material needed for the rare disease care was missing because it is now used for patients affected by COVID-19 | - Yes<br>- No |
| **4** | Since the beginning of the COVID-19 pandemic, have you/has the person you care for experienced an interruption in the following aspects of the care provided by healthcare professionals for the rare disease? | a) Rehabilitation therapy (physiotherapy, ergotherapy, speech, physical therapy, massage, etc.)<br>b) Appointment with the GP/specialist who provides care for the rare disease<br>c) Diagnosis test (lab test such as blood tests, bacteriological test, urine analysis, medical imaging, cardiac and respiratory tests, etc.)<br>d) Medical therapies at home or at the hospital (infusions, chemotherapy, hormonal treatment, etc.)<br>e) Psychiatry follow-up<br>f) Surgery or transplant | - Yes, it was completely cancelled<br>- Yes, it was postponed or delayed<br>- No |
| **5** | Since the COVID-19 pandemic started, has one of the medicines/treatments needed for the rare disease been unavailable when visiting the pharmacy or the hospital? | | - No<br>- Yes, temporarily<br>- Yes, you had to stop taking it or take an alternative |
| **6** | Do you need the following support and do you still have access to them since the COVID-19 pandemic started? | a) Family, friends or neighbours support<br>b) Psychological support<br>c) Home care (nurse, personal assistant for self-care etc.)<br>d) Social worker support<br>e) Support for house chores and daily tasks | - Yes, I need this support and I can still access it<br>- Yes, I need this support but I receive less support since the pandemic started<br>- Yes, I need this support but it stopped completely since the pandemic started<br>- I need this support but have not received it before or during the pandemic<br>- No, I don't need this support |
| **7** | How do you experience the interruptions related to the COVID-19 pandemic? | a) Detrimental to you/her/his well-being<br>b) Detrimental to you/her/his health<br>c) Life threatening | - Definitely<br>- Probably<br>- Probably not<br>- Definitely not<br>- I don't know |
| **8** | Are you afraid medicine/treatment shortages might happen in the future if the pandemic continues? | | - Yes, a little<br>- Yes, a lot<br>- No |
| **9** | With regard to COVID-19 and in relation to the rare disease, do you feel that you have access to all the information you need? | | - Most of the time<br>- Some of the time<br>- Seldom<br>- Never |
| **10** | Since the beginning of the COVID-19 pandemic and in relation to the rare disease, what are your experiences with the following forms of telemedicine? | a) Online consultations or any other form of telemedicine online or via phone<br>b) Prescription via email | - Yes, and this was new to me<br>- Yes, and it was already the case before the COVID-19 pandemic<br>- No |

| | | | |
|---|---|---|---|
| | | c) Online education and training to help you manage the rare disease yourself | |
| **11** | How would you qualify your experience with these forms of telemedicine? | | - Very useful<br>- Fairly useful<br>- Not very useful<br>- Not useful at all |
| **12** | What is your current occupation? | | - Employed<br>- Homemaker<br>- Leave of absence<br>- Retired<br>- Self-employed<br>- Student<br>- Unable to work<br>- Unemployed |
| **13** | In order to adapt to the COVID-19 pandemic, which of the following measures did your employer allow? | a) Possibility to work from home<br>b) More flexible working hours<br>c) Reduction of number of working hours<br>d) Paid carer leave | - Yes, and this was already imposed by public authorities<br>- Yes, and this was advised by public authorities<br>- Yes, and this is an initiative from my employer<br>- No |
| **14** | Please choose the sentence that best describes your situation since the COVID-19 pandemic started: | | - I work from home<br>- I had to stop working<br>- I continue to work normally<br>- I had to significantly reduce my number of working hours<br>- I am on a leave of absence<br>- I started to work part-time |
| **15** | What level of threat do you think the coronavirus poses to you/to the person with a rare disease you care for? | | - Very low threat<br>- Low threat<br>- High threat<br>- Very high threat |
| **16** | Since the COVID-19 pandemic started, how often did you feel the following: | a) you could not overcome your problems?<br>b) tensions between family members?<br>c) isolated?<br>d) unhappy and/or depressed?<br>e) strengthening of the family unit? | - Most of the time<br>- Sometimes<br>- Rarely |

*Table S2. Respondent geographic origins*

| Country | Geographic situation* | Number of respondents | Total number of patients per county§ |
|---|---|---|---|
| Albania | EEC | 1 | 123 |
| Bulgaria | EEC | 1 | 155 |
| Croatia | EEC | 11 | 93 |
| Czech Republic | EEC | 8 | 619 |
| Greece | EEC | 78 | 621 |
| Hungary | EEC | 5 | 507 |
| Latvia | EEC | 3 | 41 |
| Montenegro | EEC | 1 | 32 |
| Poland | EEC | 14 | 721 |
| Romania | EEC | 7 | 167 |
| Russia | EEC | 3 | 3269 |
| Serbia | EEC | 27 | 196 |
| Slovakia | EEC | 11 | 294 |
| Slovenia | EEC | 3 | 112 |
| Austria | WEC | 9 | 800 |
| Belgium | WEC | 34 | 1319 |
| Denmark | WEC | 26 | 510 |
| Finland | WEC | 7 | 104 |
| France | WEC | 334 | 6940 |
| Germany | WEC | 50 | 6119 |
| Italy | WEC | 17 | 5565 |
| Luxembourg | WEC | 1 | 36 |
| Netherlands | WEC | 80 | 1473 |
| Norway | WEC | 1 | 254 |
| Portugal | WEC | 10 | 341 |
| Spain | WEC | 52 | 2075 |
| Sweden | WEC | 52 | 686 |
| Switzerland | WEC | 43 | 963 |
| United Kingdom | WEC | 8 | 10468 |

* EEC = Eastern European Countries; WEC = Western European Countries

§ Numbers taken from the ECFS-Patient Registry report of 2018

*Table S3. Responses to the survey per question and option.*

| **Q1** | **Are the following measures currently in place in your country/region/state in to fight against the COVID-19 pandemic?** | | | | | | | | | | | | | | | | | |
|---|---|---|---|---|---|---|---|---|---|---|---|---|---|---|---|---|---|---|
| *Options* | *a) Social distancing, which includes avoiding mass gatherings, and maintaining distance from others when possible* | | | | | | *b) Closure of educational facilities* | | | | | | *c) Lockdown measures* | | | | | |
| Responses | Yes | | No | | I don't know | | Yes | | No | | I don't know | | Yes | | No | | I don't know | |
| WEC | 718 | 99% | 5 | 1% | 1 | 0% | 665 | 92% | 49 | 7% | 10 | 1% | 591 | 82% | 122 | 17% | 11 | 2% |
| EEC | 167 | 97% | 5 | 3% | 1 | 1% | 161 | 93% | 11 | 6% | 1 | 1% | 124 | 72% | 48 | 28% | 1 | 1% |
| TOTAL | 885 | 99% | 10 | 1% | 2 | 0% | 826 | 92% | 60 | 7% | 11 | 1% | 715 | 80% | 170 | 19% | 12 | 1% |

| **Q2** | **Are protective equipment such as face masks and plastic gloves available in your country/region/state?** | | | | | | | | | | | | | | | |
|---|---|---|---|---|---|---|---|---|---|---|---|---|---|---|---|---|
| *Options* | *a) For you* | | | | | | | | *b) For the person you care for* | | | | | | | |
| Responses | Available and easily accessible | | Difficult to find, sometimes not available | | Impossible to find, not available | | Not needed | | Available and easily accessible | | Difficult to find, sometimes not available | | Impossible to find, not available | | Not needed | |
| WEC | 89 | 13% | 313 | 45% | 256 | 37% | 43 | 6% | 35 | 11% | 140 | 45% | 111 | 36% | 25 | 8% |
| EEC | 75 | 44% | 87 | 51% | 8 | 5% | 2 | 1% | 42 | 47% | 40 | 44% | 6 | 7% | 2 | 2% |
| TOTAL | 164 | 19% | 400 | 46% | 264 | 30% | 45 | 5% | 77 | 19% | 180 | 45% | 117 | 29% | 27 | 7% |
| *Options* | *c) For your health professionals* | | | | | | | | *d) For your social care professionals* | | | | | | | |
| Responses | Available and easily accessible | | Difficult to find, sometimes not available | | Impossible to find, not available | | Not needed | | Available and easily accessible | | Difficult to find, sometimes not available | | Impossible to find, not available | | Not needed | |
| WEC | 182 | 29% | 382 | 61% | 63 | 10% | 4 | 1% | 57 | 16% | 219 | 62% | 61 | 17% | 19 | 5% |
| EEC | 80 | 47% | 89 | 53% | 0 | 0% | 0 | 0% | 57 | 36% | 95 | 61% | 4 | 3% | 1 | 1% |
| TOTAL | 262 | 33% | 471 | 59% | 63 | 8% | 4 | 1% | 114 | 22% | 314 | 61% | 65 | 13% | 20 | 4% |

| **Q3** | **Since the COVID-19 pandemic started, how was hospital care impacted for you/ for the person you care for?** | | | | | | | | | | | | | | | | | | | |
|---|---|---|---|---|---|---|---|---|---|---|---|---|---|---|---|---|---|---|---|---|
| *Options* | *a) You did not go to the hospital because you are fearful of catching COVID-19* | | | | *b) You did not go to the hospital because you are fearful that the person you care for might catch COVID-19* | | | | *c) Being told not to go to the hospital if you or the person you care for affected by a rare disease becomes unwell for other reasons than COVID-19* | | | | *d) The hospital/unit that provides care for the rare disease is closed* | | | | *e) The necessary material needed for the rare disease care was missing because it is now used for patients affected by COVID-19* | | | |
| Responses | Yes | | No | | Yes | | No | | Yes | | No | | Yes | | No | | Yes | | No | |
| WEC | 181 | 62% | 112 | 38% | 125 | 52% | 116 | 48% | 187 | 35% | 347 | 65% | 84 | 16% | 452 | 84% | 88 | 16% | 448 | 84% |
| EEC | 35 | 85% | 6 | 15% | 25 | 74% | 9 | 26% | 25 | 32% | 52 | 68% | 24 | 31% | 53 | 69% | 8 | 10% | 69 | 90% |
| TOTAL | 216 | 65% | 118 | 35% | 150 | 55% | 125 | 45% | 212 | 35% | 399 | 65% | 108 | 18% | 505 | 82% | 96 | 16% | 517 | 84% |

| **Q4** | **Since the beginning of the COVID-19 pandemic, have you/has the person you care for experienced an interruption in the following aspects of the care provided by healthcare professionals for the rare disease?** | | | | | | | | | | | | | | | | | |
|---|---|---|---|---|---|---|---|---|---|---|---|---|---|---|---|---|---|---|
| *Options* | *a) Rehabilitation therapy (physiotherapy, ergotherapy, speech, physical therapy, massage, etc.)* | | | | | | *b) Appointment with the GP/specialist who provides care for the rare disease* | | | | | | *c) Diagnosis test (lab test such as blood tests, bacteriological test, urine analysis, medical imaging, cardiac and respiratory tests, etc.)* | | | | | |
| Responses | Cancelled | | Postponed/ delayed | | No | | Cancelled | | Postponed/ delayed | | No | | Cancelled | | Postponed/ delayed | | No | |
| WEC | 318 | 56% | 135 | 24% | 114 | 20% | 101 | 16% | 349 | 55% | 184 | 29% | 96 | 17% | 265 | 48% | 196 | 35% |
| EEC | 66 | 46% | 40 | 28% | 36 | 25% | 55 | 36% | 69 | 45% | 30 | 19% | 41 | 28% | 71 | 48% | 36 | 24% |

| | | | | | | | | | | | | | | | | | | |
|---|---|---|---|---|---|---|---|---|---|---|---|---|---|---|---|---|---|---|
| TOTAL | 384 | 54% | 175 | 25% | 150 | 21% | 156 | 20% | 418 | 53% | 214 | 27% | 137 | 19% | 336 | 48% | 232 | 33% |
| *Options* | *d) Medical therapies at home or at the hospital (infusions, chemotherapy, hormonal treatment, etc.)* | | | | | | *e) Psychiatry follow-up* | | | | | | *f) Surgery or transplant* | | | | | |
| Responses | Cancelled | | Postponed/ delayed | | No | | Cancelled | | Postponed/ delayed | | No | | Cancelled | | Postponed/ delayed | | No | |
| WEC | 77 | 19% | 142 | 34% | 197 | 47% | 29 | 17% | 47 | 28% | 91 | 54% | 25 | 17% | 55 | 36% | 71 | 47% |
| EEC | 29 | 23% | 52 | 41% | 47 | 37% | 13 | 17% | 14 | 18% | 50 | 65% | 13 | 17% | 22 | 28% | 43 | 55% |
| TOTAL | 106 | 19% | 194 | 36% | 244 | 45% | 42 | 17% | 61 | 25% | 141 | 58% | 38 | 17% | 77 | 34% | 114 | 50% |

| **Q5** | **Since the COVID-19 pandemic started, has one of the medicines/treatments needed for the rare disease been unavailable when visiting the pharmacy or the hospital?** | | | | | |
|---|---|---|---|---|---|---|
| Responses | Yes, you had to stop taking it or take an alternative | | Yes, temporarily | | No | |
| WEC | 46 | 6% | 106 | 15% | 572 | 79% |
| EEC | 9 | 5% | 53 | 31% | 111 | 64% |
| TOTAL | 55 | 6% | 159 | 18% | 683 | 76% |

| **Q6** | **Do you need the following support and do you still have access to them since the COVID-19 pandemic started?** | | | | | | | | | | | | | | | | | | | |
|---|---|---|---|---|---|---|---|---|---|---|---|---|---|---|---|---|---|---|---|---|
| *Options* | *a) Family, friends or neighbours support* | | | | | | | | | | *b) Psychological support* | | | | | | | | | |
| Responses | Yes, I need this support and I can still access it | | Yes, I need this support but I receive less support since the pandemic started | | Yes, I need this support but it stopped completely since the pandemic started | | I need this support but have not received it before or during the pandemic | | No, I don't need this support | | Yes, I need this support and I can still access it | | Yes, I need this support but I receive less support since the pandemic started | | Yes, I need this support but it stopped completely since the pandemic started | | I need this support but have not received it before or during the pandemic | | No, I don't need this support | |
| WEC | 243 | 37% | 60 | 9% | 67 | 10% | 14 | 2% | 268 | 41% | 78 | 12% | 31 | 5% | 24 | 4% | 33 | 5% | 504 | 75% |
| EEC | 79 | 53% | 8 | 5% | 14 | 9% | 13 | 9% | 34 | 23% | 13 | 8% | 4 | 3% | 4 | 3% | 33 | 21% | 100 | 65% |
| TOTAL | 322 | 40% | 68 | 9% | 81 | 10% | 27 | 3% | 302 | 38% | 91 | 11% | 35 | 4% | 28 | 3% | 66 | 8% | 604 | 73% |
| *Options* | *c) Home care (nurse, personal assistant for self-care etc.)* | | | | | | | | | | *d) Social worker support* | | | | | | | | | |
| Responses | Yes, I need this support and I can still access to it | | Yes, I need this support but I receive less support since the pandemic started | | Yes, I need this support but it stopped completely since the pandemic started | | I need this support but have not received it before or during the pandemic | | No, I don't need this support | | Yes, I need this support and I can still access it | | Yes, I need this support but I receive less support since the pandemic started | | Yes, I need this support but it stopped completely since the pandemic started | | I need this support but have not received it before or during the pandemic | | No, I don't need this support | |
| WEC | 87 | 13% | 37 | 6% | 42 | 6% | 22 | 3% | 477 | 72% | 69 | 10% | 18 | 3% | 5 | 1% | 30 | 4% | 590 | 83% |
| EEC | 4 | 3% | 2 | 1% | 4 | 3% | 20 | 13% | 120 | 80% | 9 | 5% | 1 | 1% | 2 | 1% | 27 | 16% | 129 | 77% |
| TOTAL | 91 | 11% | 39 | 5% | 46 | 6% | 42 | 5% | 597 | 73% | 78 | 9% | 19 | 2% | 7 | 1% | 57 | 6% | 719 | 82% |
| *Options* | *e) Support for house chores and daily tasks* | | | | | | | | | | | | | | | | | | | |
| Responses | Yes, I need this support and I can still access it | | Yes, I need this support but I receive less support since the pandemic started | | Yes, I need this support but it stopped completely since the pandemic started | | I need this support but have not received it before or during the pandemic | | No, I don't need this support | | | | | | | | | | | |
| WEC | 18 | 3% | 15 | 2% | 47 | 7% | 45 | 7% | 537 | 81% | | | | | | | | | | |
| EEC | 16 | 11% | 0 | 0% | 5 | 3% | 19 | 13% | 109 | 73% | | | | | | | | | | |
| TOTAL | 34 | 4% | 15 | 2% | 52 | 6% | 64 | 8% | 646 | 80% | | | | | | | | | | |

| **Q7** | **How do you experience the interruptions related to the COVID-19 pandemic?** | | | | | | | | | |
|---|---|---|---|---|---|---|---|---|---|---|
| *Options* | *a) Detrimental to you/her/his well-being* | | | | | | | | | |
| Responses | Definitely | | Probably | | Probably not | | Definitely not | | I don't know | |
| WEC | 134 | 21% | 251 | 40% | 155 | 25% | 73 | 12% | 13 | 2% |
| EEC | 40 | 27% | 68 | 46% | 26 | 18% | 14 | 9% | 0 | 0% |
| TOTAL | 174 | 22% | 319 | 41% | 181 | 23% | 87 | 11% | 13 | 2% |
| *Options* | *b) Detrimental to you/her/his health* | | | | | | | | | |
| Responses | Definitely | | Probably | | Probably not | | Definitely not | | I don't know | |
| WEC | 114 | 18% | 240 | 38% | 207 | 33% | 53 | 8% | 12 | 2% |
| EEC | 37 | 25% | 73 | 48% | 28 | 19% | 10 | 7% | 3 | 2% |
| TOTAL | 151 | 19% | 313 | 40% | 235 | 30% | 63 | 8% | 15 | 2% |
| *Options* | *c) Life threatening* | | | | | | | | | |
| Responses | Definitely | | Probably | | Probably not | | Definitely not | | I don't know | |
| WEC | 47 | 8% | 141 | 23% | 257 | 41% | 157 | 25% | 24 | 4% |
| EEC | 28 | 19% | 57 | 38% | 42 | 28% | 15 | 10% | 9 | 6% |
| TOTAL | 75 | 10% | 198 | 25% | 299 | 38% | 172 | 22% | 33 | 4% |

| **Q8** | **Are you afraid medicine/treatment shortages might happen in the future if the pandemic continues?** | | | | | |
|---|---|---|---|---|---|---|
| Responses | Yes, a little | | Yes, a lot | | No | |
| WEC | 316 | 55% | 168 | 29% | 88 | 15% |
| EEC | 55 | 50% | 39 | 35% | 17 | 15% |
| TOTAL | 371 | 54% | 207 | 30% | 105 | 15% |

| **Q9** | **With regard to COVID-19 and in relation to the rare disease, do you feel that you have access to all the information you need?** | | | | | | | | | | | | | | | |
|---|---|---|---|---|---|---|---|---|---|---|---|---|---|---|---|---|
| *Age category* | *Below 35* | | | | | | | | *35 to 49* | | | | | | | |
| Responses | Most of the time | | Some of the time | | Seldom | | Never | | Most of the time | | Some of the time | | Seldom | | Never | |
| WEC | 126 | 37% | 111 | 33% | 91 | 27% | 10 | 3% | 127 | 41% | 98 | 32% | 72 | 23% | 10 | 3% |
| EEC | 34 | 45% | 24 | 32% | 13 | 17% | 4 | 5% | 35 | 45% | 36 | 46% | 7 | 9% | 0 | 0% |
| TOTAL | 160 | 39% | 135 | 33% | 104 | 25% | 14 | 3% | 162 | 42% | 134 | 35% | 79 | 21% | 10 | 3% |
| *Age category* | *50 and above* | | | | | | | | | | | | | | | |
| Responses | Most of the time | | Some of the time | | Seldom | | Never | | | | | | | | | |
| WEC | 41 | 52% | 22 | 28% | 13 | 16% | 3 | 4% | | | | | | | | |
| EEC | 11 | 55% | 7 | 35% | 2 | 10% | 0 | 0% | | | | | | | | |
| TOTAL | 52 | 53% | 29 | 29% | 15 | 15% | 3 | 3% | | | | | | | | |

| **Q10** | **Since the beginning of the COVID-19 pandemic and in relation to the rare disease, what are your experiences with the following forms of telemedicine?** | | | | | | | | | | | | | | | | | |
|---|---|---|---|---|---|---|---|---|---|---|---|---|---|---|---|---|---|---|
| *Options* | *a) Online consultations or any other form of telemedicine online or via phone* | | | | | | *b) Prescriptions via email* | | | | | | *c) Online education and training to help you manage the rare disease yourself* | | | | | |
| Responses | Yes, new to me | | Yes, same as before COVID | | No | | Yes, new to me | | Yes, same as before COVID | | No | | Yes, new to me | | Yes, same as before COVID | | No | |
| WEC | 214 | 37% | 115 | 20% | 242 | 42% | 68 | 10% | 304 | 46% | 290 | 44% | 43 | 6% | 49 | 7% | 585 | 86% |
| EEC | 56 | 38% | 31 | 21% | 62 | 42% | 52 | 34% | 45 | 30% | 55 | 36% | 14 | 9% | 19 | 12% | 130 | 80% |

| | | | | | | | | | | | | | | | | | | |
|---|---|---|---|---|---|---|---|---|---|---|---|---|---|---|---|---|---|---|
| TOTAL | 270 | 38% | 146 | 20% | 304 | 42% | 120 | 15% | 349 | 43% | 345 | 42% | 57 | 7% | 68 | 8% | 715 | 85% |

| **Q11** | **How would you qualify your experience with these forms of telemedicine?** | | | | | | | |
|---|---|---|---|---|---|---|---|---|
| Responses | Very Useful | | Fairly useful | | Not very useful | | Not useful at all | |
| WEC | 230 | 48% | 197 | 41% | 49 | 10% | 6 | 1% |
| EEC | 60 | 54% | 40 | 36% | 9 | 8% | 2 | 2% |
| TOTAL | 290 | 49% | 237 | 40% | 58 | 10% | 8 | 1% |

| **Q12** | **What is your current occupation?** | | | | | | | | | | | | | | | |
|---|---|---|---|---|---|---|---|---|---|---|---|---|---|---|---|---|
| Responses | Employed | | Homemaker | | Leave of absence | | Retired | | Self-employed | | Student | | Unable to work | | Unemployed | |
| WEC | 383 | 54% | 51 | 7% | 28 | 4% | 25 | 4% | 33 | 5% | 65 | 9% | 90 | 13% | 37 | 5% |
| EEC | 56 | 32% | 24 | 14% | 17 | 10% | 16 | 9% | 18 | 10% | 13 | 8% | 15 | 9% | 14 | 8% |
| TOTAL | 439 | 50% | 75 | 8% | 45 | 5% | 41 | 5% | 51 | 6% | 78 | 9% | 105 | 12% | 51 | 6% |

| **Q13** | **In order to adapt to the COVID-19 pandemic, which of the following measures did your employer allow?** | | | | | | | | | | | | | | | |
|---|---|---|---|---|---|---|---|---|---|---|---|---|---|---|---|---|
| *Options* | *a) Possibility to work from home* | | | | | | | | *b) More flexible working hours* | | | | | | | |
| Responses | Initiative from employer | | Advised by public authorities | | Imposed by public authorities | | No special measures | | Initiative from employer | | Advised by public authorities | | Imposed by public authorities | | No special measures | |
| WEC | 125 | 32% | 67 | 17% | 87 | 22% | 111 | 28% | 124 | 32% | 61 | 16% | 81 | 21% | 125 | 32% |
| EEC | 21 | 30% | 10 | 14% | 18 | 25% | 22 | 31% | 23 | 32% | 11 | 15% | 22 | 31% | 15 | 21% |
| TOTAL | 146 | 32% | 77 | 17% | 105 | 23% | 133 | 29% | 147 | 32% | 72 | 16% | 103 | 22% | 140 | 30% |
| *Options* | *c) Reduction of number of working hours* | | | | | | | | *d) Paid carer leave* | | | | | | | |
| Responses | Initiative from employer | | Advised by public authorities | | Imposed by public authorities | | No special measures | | Initiative from employer | | Advised by public authorities | | Imposed by public authorities | | No special measures | |
| WEC | 64 | 16% | 18 | 5% | 36 | 9% | 272 | 70% | 22 | 6% | 5 | 1% | 24 | 6% | 339 | 87% |
| EEC | 15 | 21% | 7 | 10% | 12 | 17% | 37 | 52% | 8 | 11% | 3 | 4% | 14 | 20% | 45 | 64% |
| TOTAL | 79 | 17% | 25 | 5% | 48 | 10% | 309 | 67% | 30 | 7% | 8 | 2% | 38 | 8% | 384 | 83% |

| **Q14** | **Please choose the sentence that best describes your situation since the COVID-19 pandemic started:** | | | | | | | | | | | |
|---|---|---|---|---|---|---|---|---|---|---|---|---|
| Responses | Work from home | | Stopped working | | Normal situation | | Reduced hours | | Leave of absence | | Started working part-time | |
| WEC | 217 | 52% | 124 | 30% | 36 | 9% | 24 | 6% | 10 | 2% | 5 | 1% |
| EEC | 41 | 46% | 23 | 26% | 13 | 15% | 8 | 9% | 4 | 4% | 0 | 0% |
| TOTAL | 258 | 51% | 147 | 29% | 49 | 10% | 32 | 6% | 14 | 3% | 5 | 1% |

| **Q15** | **What level of threat do you think the coronavirus poses to you/to the person with a rare disease you care for?** | | | | | | | |
|---|---|---|---|---|---|---|---|---|
| Responses | Very low threat | | Low threat | | High threat | | Very high threat | |
| WEC | 4 | 1% | 59 | 8% | 264 | 37% | 389 | 54% |
| EEC | 1 | 1% | 8 | 5% | 55 | 33% | 102 | 61% |

| | | | | | | | | |
|---|---|---|---|---|---|---|---|---|
| TOTAL | 5 | 1% | 67 | 8% | 319 | 36% | 491 | 56% |

| Q16 | **Since the COVID-19 pandemic started, how often did you feel the following:** | | | | | | | | | | | | | | | | | |
|---|---|---|---|---|---|---|---|---|---|---|---|---|---|---|---|---|---|---|
| *Options* | *a) You could not overcome your problems?* | | | | | | *b) Tensions between family members?* | | | | | | *c) Isolated?* | | | | | |
| Responses | Rarely | | Sometimes | | Most of the time | | Rarely | | Sometimes | | Most of the time | | Rarely | | Sometimes | | Most of the time | |
| WEC | 388 | 55% | 206 | 29% | 107 | 15% | 315 | 45% | 251 | 36% | 129 | 19% | 252 | 35% | 214 | 30% | 250 | 35% |
| EEC | 83 | 51% | 55 | 34% | 26 | 16% | 79 | 48% | 66 | 40% | 21 | 13% | 52 | 31% | 49 | 29% | 66 | 40% |
| TOTAL | 471 | 54% | 261 | 30% | 133 | 15% | 394 | 46% | 317 | 37% | 150 | 17% | 304 | 34% | 263 | 30% | 316 | 36% |
| *Options* | *d) Unhappy and/or depressed?* | | | | | | *e) Strengthening of the family unit?* | | | | | | | | | | | |
| Responses | Rarely | | Sometimes | | Most of the time | | Rarely | | Sometimes | | Most of the time | | | | | | | |
| WEC | 294 | 41% | 254 | 36% | 165 | 23% | 95 | 14% | 186 | 27% | 414 | 60% | | | | | | |
| EEC | 62 | 35% | 65 | 37% | 50 | 28% | 21 | 13% | 34 | 21% | 106 | 66% | | | | | | |
| TOTAL | 356 | 40% | 319 | 36% | 215 | 24% | 116 | 14% | 220 | 26% | 520 | 61% | | | | | | |

*Table S4. List of variables and attributed quantitative values that were used for to calculate correlations between psychological status and potentially impacting factors.*

| | | | | |
|---|---|---|---|---|
| **Psychosocial status** | | | | |
| ***Q*** | ***Question*** | ***Options*** | ***Responses retained*** | ***Quantitative values*** |
| **15** | What level of threat do you think the coronavirus poses?<br>a) As a PWCF<br>b) For the PWCF you care for | | - Very low threat<br>- Low threat<br>- High threat<br>- Very high threat | 0<br>1<br>2<br>3 |
| **16** | How often did you feel the following: | a) you could not overcome your problems?<br>b) tensions between family members?<br>c) isolated?<br>d) unhappy and/or depressed?<br>e) strengthening of the family unit? | - Rarely<br>- Sometimes<br>- Most of the time | 0<br>1<br>2 |
| **Potentially impacting factors** | | | | |
| ***Q*** | ***Question*** | ***Options*** | ***Responses retained*** | ***Quantitative values*** |
| **1** | Are the following measures currently in place? | a) Social distancing<br>b) Closure of educational facilities<br>c) Lockdown measures | - No<br>- Yes | 0<br>1 |
| **2** | Are protective equipment available? | a) For you<br>b) For the person you care for<br>c) For your health professionals<br>d) For your social care professionals | - Available<br>- Sometimes not available<br>- Not available | 0<br>1<br>2 |
| **4** | Have you experienced an interruption in your care? | a) Rehabilitation therapy<br>b) Appointments with GP or specialist<br>c) Diagnosis test<br>d) Medical therapies at home or at the hospital<br>e) Psychiatry follow-up<br>f) Surgery or transplant | - No<br>- Postponed/delayed<br>- Cancelled | 0<br>1<br>2 |
| **5** | Has one of your medicines/treatments been unavailable? | | - No<br>- Temporarily<br>- Yes | 0<br>1<br>2 |
| **6** | Do you still have access to the following support? | a) Family, friends or neighbours support<br>b) Psychological support<br>c) Home care<br>d) Social worker support<br>e) Support for house chores and daily tasks | - I can still access it<br>- I receive less support<br>- It stopped | 0<br>1<br>2 |
| **7** | How do you experience the interruptions related to the COVID-19 pandemic? | a) Detrimental to you/her/his well-being<br>b) Detrimental to you/her/his health<br>c) Life threatening | - Definitely not<br>- Probably not<br>- Probably<br>- Definitely | 0<br>1<br>2<br>3 |
| **8** | Are you afraid medicine/treatment shortages might happen in the future if the pandemic continues? | | - No<br>- A little<br>- A lot | 0<br>1<br>2 |
| **9** | Do you feel that you have access to all the information you need? | | - Never<br>- Seldom<br>- Some of the time<br>- Most of the time | 0<br>1<br>2<br>3 |
| **15** | What level of threat do you think the coronavirus poses?<br>a) As a PWCF<br>b) For the PWCF you care for | | - Very low threat<br>- Low threat<br>- High threat<br>- Very high threat | 0<br>1<br>2<br>3 |
| **16** | How often did you feel the following: | a) you could not overcome your problems?<br>b) tensions between family members?<br>c) isolated?<br>d) unhappy and/or depressed?<br>e) strengthening of the family unit? | - Rarely<br>- Sometimes<br>- Most of the time | 0<br>1<br>2 |

*Table S5. Respondent characteristics*

| Numbers of respondents | | WEC* | EEC* | Total |
|---|---|---|---|---|
| **Gender** | Females | 565 (78%) | 137 (79%) | 702 (78%) |
| | Males | 157 (22%) | 36 (21%) | 193 (22%) |
| | Other | 2 (0,2%) | 0 (0%) | 2 (0,2%) |
| **Respondents** | Patients | 394 (54%) | 75 (43%) | 469 (52%) |
| | Others | 330 (46%) | 98 (57%) | 428 (48%) |
| **Age** | Below 35 | 338 (47%) | 75 (43%) | 413 (46%) |
| | 35 to 49 years old | 307 (42%) | 78 (45%) | 385 (43%) |
| | 50 and above | 79 (11%) | 20 (12%) | 99 (11%) |

* EEC = Eastern European Countries; WEC = Western European Countries

*Figure S1. Numbers of respondents according to age, gender, for PWCF and others, in WEC and EEC.*

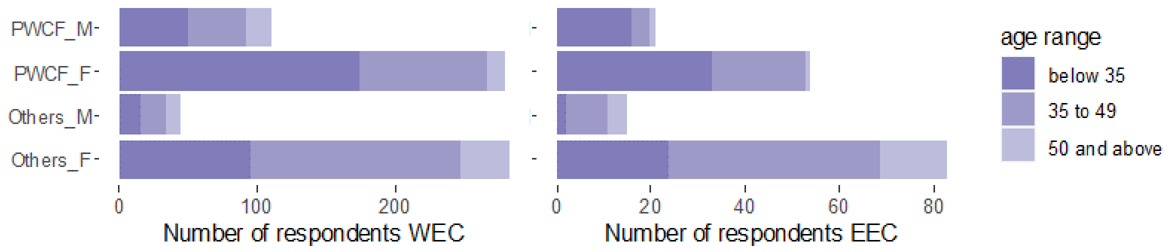


*Figure S2. Theme 1: Country situation and availability of protective equipment. Distribution of the responses to Q2 expressed as percentages of the total number of respondents.*

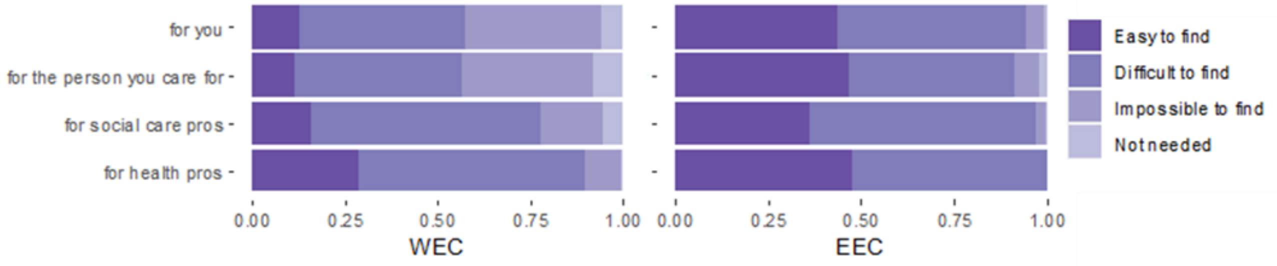

*Table S6. Summary of significant correlations calculated between responses to Q15 and Q16 on one hand, and Q1, Q2, Q4, Q5, Q6, Q7, Q8, Q9, Q15 and Q16 on the other hand, using the numerical values reported in Table S4.*

Correlation coefficients (CC) are shown only when the associated *p*-value was below 1,9E-04; CC are gradually coloured from -1 (red) to green (+1).

| | | Q15a. Level of threat (as PWCF) | | Q15b. Level of threat (for the PWCF cared for) | | Q16a. Difficulty to overcome problems | | Q16b. Family tensions | | Q16c. Feeling of isolation | | Q16D. Feeling depressed | | Q16e. Family strengthening | |
|---|---|---|---|---|---|---|---|---|---|---|---|---|---|---|---|
| | | CC | *p*-value | CC | *p*-value | CC | *p*-value | CC | *p*-value | CC | *p*-value | CC | *p*-value | CC | *p*-value |
| Q4b | Interruptions in follow-up appointments | | | | | 0,15 | 1,2E-05 | | | 0,13 | 3,4E-05 | | | | |
| Q5 | Interruptions in treatment | | | | | 0,13 | 8,6E-05 | | | | | 0,14 | 3,8E-05 | | |
| Q6a | Less lamily support | | | | | | | | | 0,19 | 1,1E-05 | | | | |
| Q7a | Detrimental to wellbeing | | | 0,27 | 7,8E-06 | 0,24 | 1,3E-12 | | | 0,15 | 4,9E-06 | 0,24 | 1,6E-13 | | |
| Q7b | Detrimental to health | 0,26 | 9,9E-06 | 0,30 | 6,0E-07 | 0,15 | 6,4E-06 | | | | | 0,15 | 1,3E-05 | | |
| Q7C | Life threatening | 0,26 | 9,9E-06 | 0,30 | 6,0E-07 | 0,15 | 6,4E-06 | | | | | 0,15 | 1,3E-05 | | |
| Q8 | Fear of shortages | | | 0,25 | 6,0E-05 | 0,16 | 2,6E-05 | | | | | | | | |
| Q9 | Sufficient information | | | | | -0,14 | 4,3E-06 | | | | | | | | |
| Q16a | Difficulty to overcome problems | | | | | | | 0,27 | 1,8E-18 | 0,38 | 1,9E-37 | 0,56 | 6,0E-77 | | |
| Q16b | Family tensions | | | | | 0,27 | 9,6E-19 | | | 0,23 | 1,3E-14 | 0,33 | 2,6E-27 | -0,21 | 2,8E-11 |
| Q16c | Feeling of isolation | | | | | 0,38 | 5,5E-36 | 0,23 | 7,9E-14 | | | 0,46 | 1,3E-54 | -0,15 | 1,5E-06 |
| Q16d | Feeling depressed | | | | | 0,56 | 3,2E-77 | 0,33 | 5,5E-27 | 0,46 | 3,8E-57 | | | -0,17 | 5,4E-08 |
| Q16e | Family strengthening | | | | | | | -0,21 | 3,1E-11 | -0,15 | 7,8E-07 | -0,17 | 5,0E-08 | | |
